# The intergenerational effects of parental physical activity on offspring brain and neurocognition in humans: a scoping review

**DOI:** 10.1101/2022.09.12.22279883

**Authors:** Sarah R. Valkenborghs, Paige C. Dent, Chelsea M. Stillman

## Abstract

Animal models suggest physical activity (PA) has intergenerational effects on brain health and neurocognition. This scoping review compiles the human literature in this area, identifies knowledge gaps, and makes recommendations for future research.

We systematically searched for experimental or observational studies conducted in humans, published in English, and reporting parental PA exposure (preconception or prenatal) and subsequent offspring brain and neurocognition. Two reviewers independently screened studies according to predetermined inclusion criteria.

Fourteen articles were included (four experimental and 10 observational). All studies reported maternal characteristics, whereas only one (7%) study reported paternal characteristics (but not paternal PA). Prenatal maternal PA exposure was examined in 10 (71%) studies, while preconception *and* prenatal PA exposure was examined in four (29%) studies. Maternal PA exposure was positively related to offspring brain and neurocognitive development in most studies.

Little is known about the intergenerational effects of parental PA on offspring brain and neurocognition in humans, particularly paternal preconception PA. More experimental studies with longer offspring follow-up and more objective and/or mechanistic assessments are required.

**HIGHLIGHTS:**

- Animal models suggest physical activity has intergenerational neurobiological effects
- All observational human studies report a positive relationship between maternal physical activity and offspring brain and neurocognition
- Most experimental human studies report no effects of maternal physical activity on offspring brain and neurocognition

## 1. INTRODUCTION

Maternal preconception and prenatal health behaviours such as alcohol consumption, diet, and smoking are known to impact on offspring neurodevelopment ^1-7^. Over the past three decades, public health interventions aimed at modifying health behaviours of pregnant women have proven largely successful ^8-11^. Physical activity (PA) is important during pregnancy as it has been linked to improved health outcomes in the mother during preconception, pregnancy, and postpartum and reduced risk of pregnancy complications ^12-14^. While pregnancy has been documented as a ‘teachable moment’ during which women have increased motivation to undertake healthy lifestyle behaviours, it is usually associated with decreased participation in PA^14-16^. Sadly, only 3 in 10 pregnant women meet PA guidelines ^17-19^. While concerns about safety and potential adverse effects on the developing fetus is one of the most frequently reported barriers to PA participation during pregnancy ^14,20^, there is no evidence of detrimental effects of PA on fetal development, risk of miscarriage, or preterm birth ^14,19^. Knowledge of fetal health benefits has been identified as a key motivator to PA during pregnancy ^21^. A recent delphi study highlights establishing the short- and long-term fetal benefits of PA in pregnancy is one of the top 10 research priorities identified by pregnant women and health care/exercise professionals ^22^. Taken together, studies that elucidate the link between PA and fetal health may prove to be more effective in motivating women to remain physically active during pregnancy.

PA not only helps to improve physical health but can also enhance brain health and neurocognition across the lifespan ^23,24^. Numerous cross-sectional studies have demonstrated that people with higher cardiorespiratory fitness (CRF) or leisure-time PA levels perform better on cognitive tasks, particularly those measuring executive or memory functions ^25^. Higher CRF is also associated with better brain structural integrity (e.g., gray matter volume) and function (e.g., activation and metabolism) in brain regions that support executive and memory functions, including the hippocampus and prefrontal cortex ^26,27^. Prospective longitudinal epidemiological studies provide convergent support to this cross-sectional evidence by demonstrating that CRF confers significant protection against normal and pathological cognitive declines in aging. For example, engaging in moderate intensity PA at a frequency of three or more times per week reduced the risk of cognitive impairment or dementia five years later ^28^. Furthermore, aerobic activities in young adulthood not only predict better cognitive performance in young adulthood ^29^, but also better cognitive and brain health decades later ^30^. Finally, recent reviews confirm that PA (especially interventions lasting six months or longer) consistently improves memory and executive functioning, as well as brain structure and function in specific brain regions ^25,31^. In sum, there is converging evidence that PA enhances neuroplasticity and inhibits neurodegenerative processes, resulting in better brain health and function across the lifespan.

Accumulating evidence in animal models suggests that PA can also have intergenerational effects on brain health and development. That is, parental PA levels—both preconception and during pregnancy—can affect the offspring brain. A number of recent meta-analyses report that parental PA enhances offspring brain growth factor expression (i.e., brain derived-neurotrophic factor, tropomyosin receptor kinase B, vascular endothelial growth factor), brain structure (neurogenesis), and neurobehaviour (learning, memory, and anxiety) ^32,33^. Larger effects are generally observed when parental PA starts preconception, however, exercising only during pregnancy also has positive effects ^32^. Effects seem consistent regardless of whether mothers or fathers are physically active and across all offspring age groups (prenatal-adulthood), indicating that parental PA exerts long-lasting benefits on offspring brain structure and neurobehaviour ^32^.

### 1.1 Rationale and aims

Despite several recent reviews of the animal literature reporting on the intergenerational effects of parental PA on offspring brain health and neurocognition, the extent of evidence of these effects in humans is unknown. This scoping review will compile the human literature in this field with the aims of (a) collating the existing human research evidence, (b) identifying knowledge gaps, and (c) making recommendations for future human research in this area. To achieve these aims the review will systematically and comprehensively synthesise the literature according to key potential moderators including sex of parental PA exposure examined (maternal and/or paternal), timing of PA exposure (preconception, prenatal, or both), timing of offspring follow-up, as well as measures of offspring brain health and neurocognition investigated.

## 2. METHODS

The conduct and reporting of this review adheres to guidelines for scoping reviews ^34,35^. We conducted a systematic literature search of PubMed, MEDLINE, Embase, Cochrane Central, and PsycINFO from database inception to 9^th^ December 2021. Our search was limited to studies conducted in humans and published in the English language. The search strategy was modified to comply with each database search criteria (Supplemental Information 1).

### 2.1 Study eligibility criteria

Studies were included if they reported data on parental PA exposure preconception (fathers and/or mothers) or prenatally (mothers only), as well as data on offspring neurocognitive development. Studies could be observational or experimental in design. Only articles published in peer-reviewed journals were included. Studies were excluded if they met any of the following criteria: (1) not published in English; (2) not human subjects; (3) published conference proceedings; or (4) theses.

### 2.2 Study Selection

After removal of duplicates, two reviewers independently screened articles in Covidence ^36^. Titles and abstracts were used to classify articles as ‘possibly relevant’ or ‘definitely irrelevant’. Articles identified as ‘definitely irrelevant’ by both reviewers were excluded. Articles identified as ‘possibly relevant’ by either reviewer progressed to full text review where they were classified as ‘include’ or ‘exclude’. Records classified as ‘include’ by both reviewers were included. Conflicts between reviewers were independently re-assessed by both reviewers. If agreement was not reached, consensus was sought through discussion. Agreement between reviewers was calculated in terms of proportionate agreement and the Cohen’s Kappa statistic ^37^.

### 2.3 Data Extraction and Synthesis

Data from the included studies were extracted by one reviewer and verified by a second reviewer. Data extraction included:

⍰ Study characteristics (e.g., design, sample size, country);
⍰ Parental characteristics (e.g., age, sex, ethnicity, education, body composition, fitness);
⍰ Offspring characteristics (e.g., age, sex, gestational age, method of delivery, body composition);
⍰ Parental PA exposure (e.g., volume, frequency, intensity, type and timing relative to pregnancy);
⍰ Offspring brain and neurocognition (e.g., measures of brain development, neurodevelopment, communication, verbal IQ, academic performance and intelligence);
⍰ Confounders controlled for in analyses; and
⍰ Summary of key findings.

Results of studies were summarised qualitatively for comparison. For the purposes of this review, these measures of intensity have been interpreted according to agreed definitions for the standardised categorisation of exercise intensity for subjective and objective measures, where applicable ^38^. Quantitative assessment was limited to tallying the number of studies reporting the same/similar outcome. Data are reported as mean (standard deviation), mean±standard error of the mean, mean [95% CI], and median {IQR}.

## 3. RESULTS

### 3.1 Flow of studies through the review

Initially 2,428 articles were identified. After removing duplicates, 2,226 articles were screened by title and abstract. Twenty-nine articles were progressed to full text review, of which 14 were included (agreement = 93%, κ = 0.86) (Figure 1) ^39-52^.

**Figure 1.**
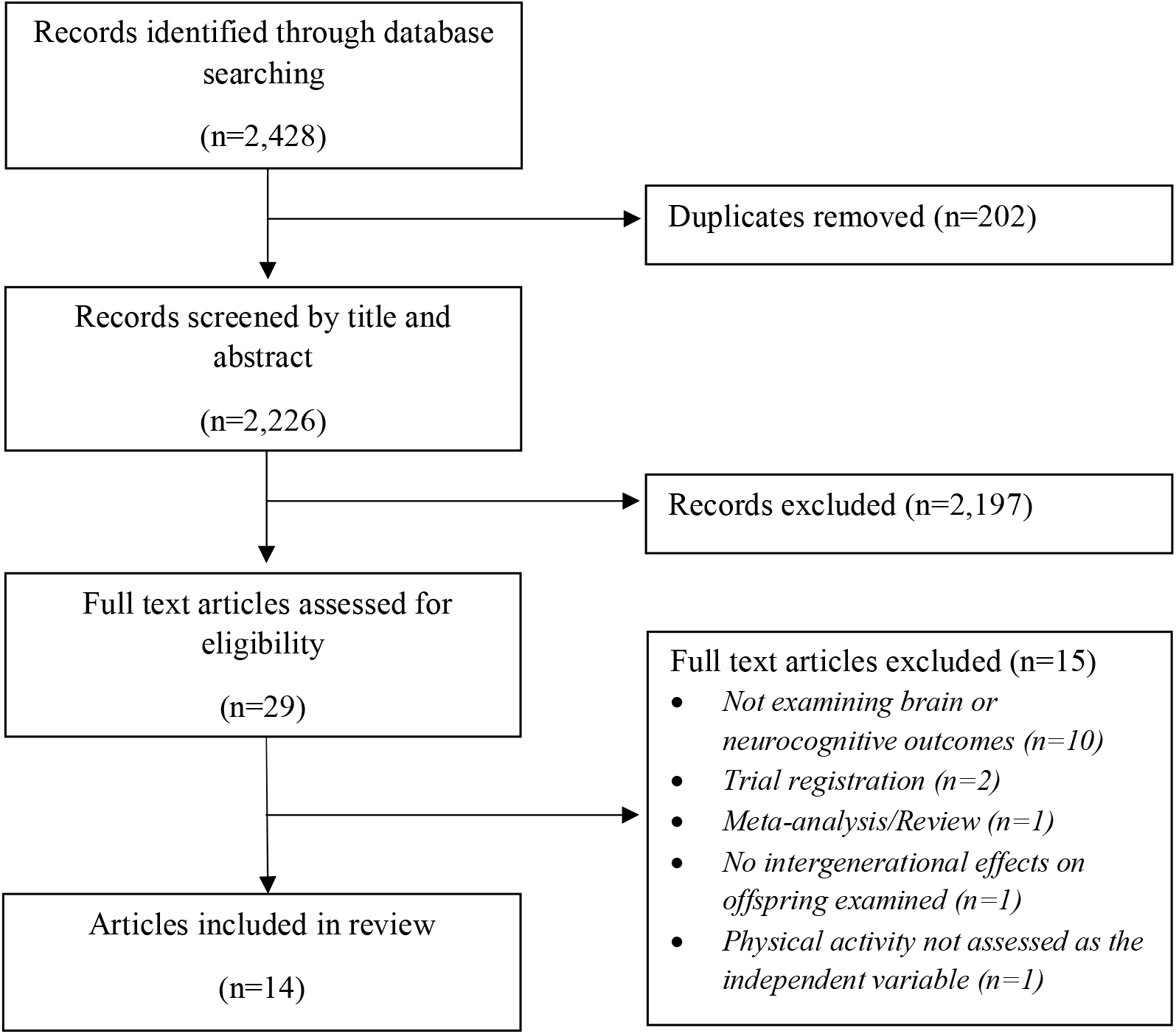
Flow of studies through the review

### 3.2 Characteristics of included studies

The publication year of included studies ranged from 1996 ^39,52^ to 2021 ^50^. Four studies (29%) were from the United States of America ^39-41,47^, two (14%) were from Norway ^43,45^, while there was one (7%) study each from Brazil ^42^, Canada ^48^, Denmark ^46^, Japan ^50^, New Zealand ^52^, Poland ^51^, and Spain ^44^. One study ^49^ reported data on two samples – one from Finland and one from the Netherlands. Four (29%) experimental studies were included (three RCTs ^43,45,48^ and one long-term follow-up of two RCTs ^49^), and with sample sizes ranging from n=18 ^48^ to n=336 ^45^. Ten (71%) observational studies were included (six prospective ^39-42,47,51^ and four retrospective ^44,46,50,52^) with the sample sizes ranging from 40 ^39^ to 74,971 ^50^.

### 3.3 Parental characteristics

All studies (100%) reported data on maternal characteristics, whereas only one (7%) study reported data on paternal age (32.5 (5.5) years) ^51^. Maternal age was reported in all but one study ^44^ and ranged from <19 years ^42^ to >35 years ^47,50^. Seven studies ^43,45,46,48-51^ reported maternal body mass index which ranged from <18.5 kg/m^2 46^ to 36 (3) kg/m^2 49^. Two studies reported maternal percentage body fat which ranged from 16.6±1.3% ^39^ to 21.0±0.8 ^41^, while pregnancy weight gain was reported in three studies ^39-41^ and ranged from 12.8±1.1 kg to 13.6±1.4 kg for participants who exercised and 16.2±1.4kg to 17.9±1.7 kg for controls / participants who did not exercise. Preconception maternal cardiorespiratory fitness (maximal oxygen consumption) was reported by two studies and ranged from 48.2±2.7 mL/kg/min ^41^ to 54.4±2.6 mL/kg/min ^39^.

### 3.4 Maternal PA exposure

#### 3.4.1 Experimental studies

The effect of maternal preconception PA alone was not examined by any experimental study, but one study reported effects of PA performed as part of a lifestyle intervention before and/or during pregnancy ^49^ (Supplemental Table 2). Otherwise, maternal preconception PA levels were reported by three RCTs and ranged from 1.0 (0-5) ^45^ to 1.7 (1.4) ^43^ exercise sessions per week, or a score of 11.0 on the Kaiser Physical Activity Survey ^48^, and were balanced across groups in all studies.

The characteristics of PA interventions (Supplemental Table 1) varied considerably between the experimental studies and were generally not reported consistently nor according to the FITT principle ^53^. Frequency was most commonly three sessions per week (three studies ^43,48,49^), but ranged from one ^45^ to five ^49^ sessions per week. Where reported, exercise consisted of a combination of aerobic, strength, and balance activities ^43^ performed at moderate ^45,48^, or moderate-vigorous^49^ intensity.

Exercise session duration ranged from ≥20 ^48^ to 60 ^43^ minutes, while the duration of the intervention was only reported by one study ^43^ and was 12 weeks between 20-36 weeks gestation. Hellenes and colleagues^45^ also delivered the intervention between 20-36 weeks gestation but did not specify the length of the program. In addition to the prescribed exercise, the two RCTs pooled by Menting et al. also asked participants to complete ≥10,000 steps per day ^49^.

#### 3.4.2 Observational studies

Maternal PA levels were most commonly retrospectively recorded subjectively by means of self-reported surveys and/or interviews in observational studies. In the three studies that examined preconception and prenatal PA participation levels ^40,44,50^, the assessment of PA varied. Nakahara et al., used responses to the International Physical Activity Questionnaire to define participants into five groups for each timepoint based on no PA (17% participants before pregnancy; 23% participants during pregnancy) or quartiles of PA which ranged from 245-8,078 METmin/week before pregnancy and 196-4774 METmin/week during pregnancy ^50^. Based upon self-reported recall 10 years post-partum, Esteban-Cornejo et al., grouped participants dichotomously according to whether they were active before (45%) and during (41%) pregnancy or not ^44^. They also created subgroups of participants who remained active (30%), became inactive (15%), became active (11%), or remained inactive (44%) during pregnancy ^44^. Finally, using weekly logbooks, one study compared a group of pregnant women who regularly performed aerobic exercise (≥55-70% VO_2max_ for ≥20min and ≥3 times per week) to a group of otherwise active pregnant women ^40^.

PA levels were examined during the prenatal period only in seven observational studies ^39,41,42,46,47,51,52^. Two studies ^39,41^ compared physically active control groups to women who continued to perform structured exercise training throughout pregnancy which consisted of aerobic activities (e.g., running, swimming, aerobics, use of ergometers, etc.) performed at moderate-vigorous ^39^or vigorous ^41^ intensity for ≥20 minutes ^41^, three ^41^ to 10 ^39^ times per week. Two studies grouped participants dichotomously according to whether they were meeting the PA guidelines or not ^42,51^. Domingues et al., administered a custom made questionnaire at birth to retrospectively determine whether participants performed at least 150 minutes of leisure time PA during the three trimesters of pregnancy (1st trimester = 7%, 2nd trimester = 5%, 3rd trimester = 4%)^42^, while Polanska et al., classified participants according to whether they met the American College of Obstetrics and Gynecology’s PA recommendation^54^ for ≥3 METs and ≥2.5 h/week (16%) or not (i.e., <3 METs or ≥3 METs and <2.5 h/week) (84%)^51^. Jukic et al., measured prenatal PA at 18 weeks gestation using the leisure time PA index as well as a survey regarding general PA (53% performed less than two hours per week)^47^. Jochumsen et al., assessed maternal PA levels using the Saltin□Grimby Physical Activity Level Scale as well as sports participation levels during the first trimester and reported that 43% were sedentary and 51% participated in light PA, while 74% did not participate in any sports^46^. Pryor et al., interviewed mothers of offspring born small-for-gestational age and asked them to retrospectively recall whether they performed little or no exercise (45%), moderate levels of exercise (45%), or high levels of exercise (11%) during pregnancy ^52^.

### 3.5 Offspring characteristics and measures of neurocognition

Offspring sex was reported in all but three studies ^45,47,48^ and was generally well-balanced (ranging from 40% ^39^ to 54% ^43^ male), except for one study which only included male offspring ^46^. Offspring follow-up was during the neonatal stage (n=2, 14%) ^40,48^, infant stage (1-2y) (n=7, 50%) ^41,42,45,47,50,51^, early childhood (4-7y) (n=3, 21%) ^39,43,49^, late childhood (8-10y) (n=2, 14%) ^44,47^, and young adulthood (17-21y) (n=1, 7%) ^46^.

The most commonly reported outcome of offspring brain and neurocognitive development was neurodevelopment which was most commonly measured with the Bayley Scales of Infant Development ^41,45,51,52^. Less common measures of neurodevelopment included the Brazelton Neonatal Behavioural Assessment Scales ^40^, Battelle’s Developmental Inventory ^42^, Five-to-Fifteen Questionnaire ^43^, and the Ages and Stages Questionnaire ^49,50^. Other outcomes of offspring brain and neurocognitive development included measures of intelligence such as the Wechsler Intelligence Scale ^39,42,47^ and the Børge Priens test score ^46^, brain activity (electroencephalography) ^48^, communication (MacArthur Infant Communication scale) ^47^, academic performance (math and language grades, grade point averages) ^44^ and behaviour (Child Behavior Checklist) ^49^.

### 3.6 Relationships between parental PA and offspring brain and neurocognitive development

All studies that examined preconception and prenatal maternal PA exposure ^40,44,50^ and eight of 11 studies that examined prenatal maternal PA exposure only ^39,41,42,46-48,51,52^ found a positive relationship with offspring brain and neurocognitive development. Maternal PA exposure was positively related to offspring brain and neurocognitive development as neonates (2/2 studies)^40,48^, infants (5/7 studies)^41,42,50-52^ and young adults (1/1 studies) ^46^ (Table 1). The majority of studies in early childhood (3/4) ^43,47,49^ did not find an association between maternal PA exposure and offspring brain and neurocognitive development, whereas the results of studies with follow up in late childhood (2/2) were mixed ^44,47^ (Supplemental Table 2). Irrespective of timing of maternal PA exposure or offspring age, while all 10 observational studies found a positive relationship between maternal PA exposure and measures of offspring brain and neurocognitive development ^39-42,44,46,47,50-52^, three of four experimental studies found no effect ^43,45,49^.

**Table 1.**
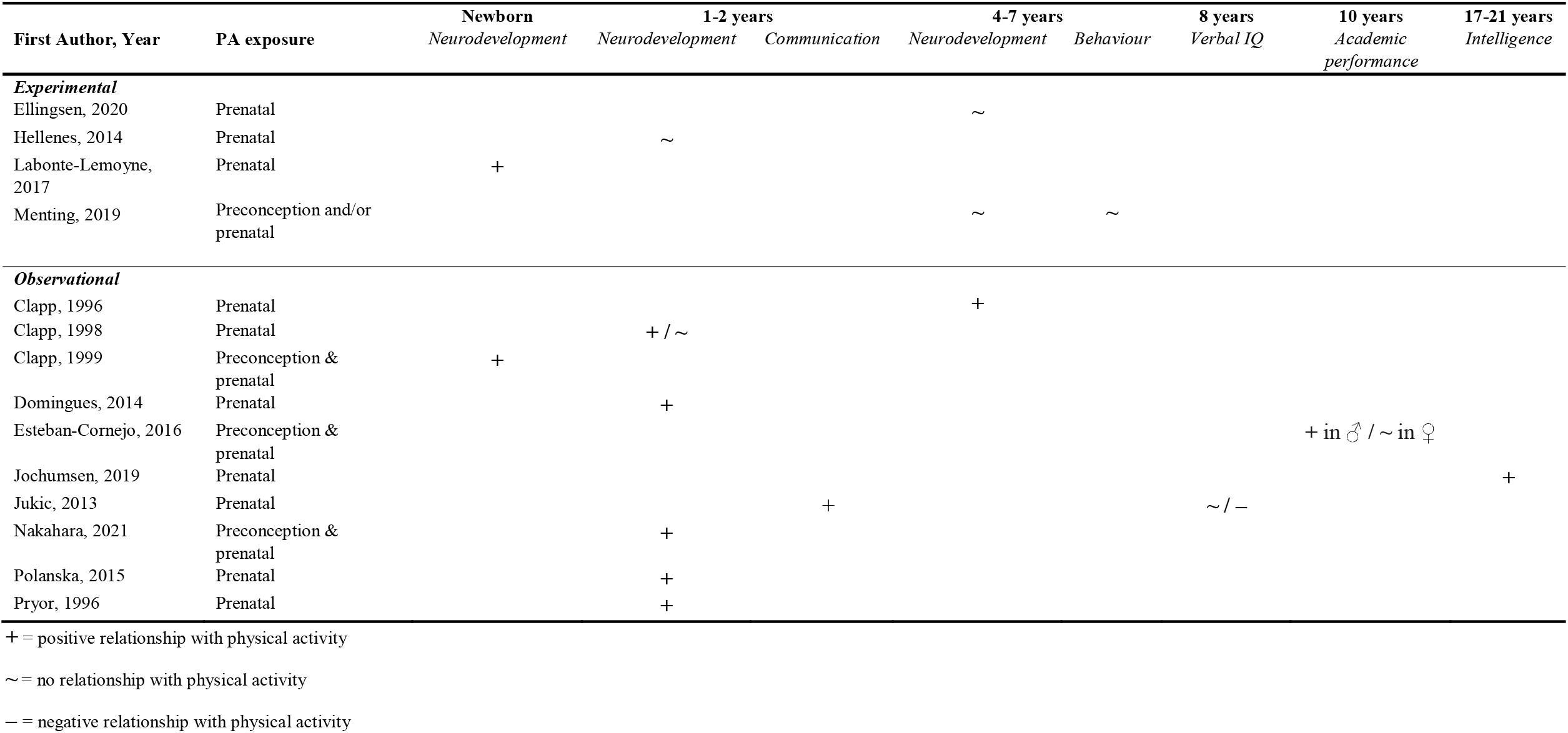
Summary of findings from included studies

## 4. DISCUSSION

In this scoping review, we systematically compiled the human literature on the intergenerational effects of parental PA on offspring brain health and neurocognition. Fourteen studies were included in the review, of which four were experimental studies focused on effects of prenatal maternal PA interventions while 10 were observational. Four studies examined preconception and prenatal maternal PA exposure and 10 examined prenatal maternal PA exposure only. While the majority of studies found that parental PA exposure was positively related to offspring brain and neurocognitive development, all studies focused on maternal PA exposure only, were mostly observational in design, and assessed infant neurodevelopment using non-objective measurement tools. Therefore, despite several recent reviews of the animal literature reporting on the beneficial intergenerational effects of parental PA on offspring brain development and neurocognition, this phenomenon is comparatively understudied in humans. Furthermore, there are several overarching limitations of the existing human evidence on this topic that present opportunities for future research including timing and method of assessment of offspring, as well as the potential mechanisms, mediators, and moderators of any effects.

### 4.1 Adherence and attrition

Characteristics of PA interventions were generally not reported, not reported consistently, or in sufficient detail to enable replication by future experimental studies and in clinical practice ^55^ nor comparison of effects of different PA programs by future systematic reviews. Therefore, future experimental studies should report intervention characteristics according to the CERT checklist^56^ or, at the very least, in the FITT format ^53^. In the studies that conducted a PA intervention, adherence to the intervention was typically not reported, not clear, or was very low. For example, one study reported that only 56% of participants adhered to the PA intervention ^43^, while another reported 64% adhered ^45^. Interestingly, neither of these studies found an effect of prenatal maternal PA on offspring neurocognitive outcomes, but this could potentially be at least partially explained by poor intervention adherence.

The same studies also reported high rates of attrition (49% at 18 months offspring follow-up ^45^ and 68% at 7 years offspring follow-up ^43^) which may have further impacted upon capacity to detect any intergenerational effects of the PA intervention (i.e., smaller than expected effect size due to poor intervention adherence combined with smaller than expected sample size due to attrition). High rates of attrition were also notable for the observational studies in this field. For example, one study experienced 50% attrition between mothers recruited and assessed during pregnancy and offspring followed up at 2 years of age ^51^. Retention of participants in this field seems disproportionately poor when compared to that typically observed in pregnancy/birth cohort studies (e.g., mean retention at follow-up ≤2 years = 87.2%; 95% CI, 81.7–92.8)) ^57^. While some attrition is unavoidable, retaining adequate participants ensures findings remain representative and unbiased. Therefore, it is imperative that future studies in this field employ a range of retention strategies tailored to both the cohort and each participant ^58-60^.

### 4.2 Assessments of neurocognition and mechanisms

While the weight of the existing human evidence clearly points to a positive or neutral effect of PA on brain health and neurocognition in newborns and infants, it is difficult to compare effects meaningfully across studies because of the heterogeneity of assessments/measures used. Further, some of the assessments utilised were developed to screen for abnormal development (e.g., ASQ) and are not suited for detecting interindividual variability and/or giftedness in neurocognitive function within otherwise typically developing infants and children, such as assessments like the Bayley Scales of Infant Development and the Brazelton Neonatal Behavioural Assessment Scales. Relatedly, it should be noted that while one study observed a relationship between prenatal maternal PA and infant mental development in offspring who were born small-for-gestational age, but not those born at an appropriate size-for-gestational-age (as measured by Bayley Scales of Infant Development), this could have been due to the considerably smaller sample size in the appropriate-sized (n=20) compared to the small-sized group (n=65) ^52^.

Additionally, only one included study used an objective and/or non-behavioural task to probe the possible neural mechanisms of intergenerational PA in humans ^48^. One direction for future studies would be to include neuroimaging outcomes (e.g., magnetic resonance imaging, functional near-infrared spectroscopy, or event-related potentials) alongside neurocognitive testing.

### 4.3 Mediators

No studies explored potential mediators of the effects of intergenerational PA on offspring brain development and neurocognition. Many of the best evidenced risk factors for adverse offspring brain and neurodevelopment are modifiable by PA, including poor maternal mental (e.g., stress, depression, anxiety), cardio-metabolic (e.g., diabetes, hypertension, obesity), and sleep health. Many of the domains of maternal mental health that are linked with aberrant offspring brain development ^61-63^, impaired neurodevelopment ^64,65^, and cognition ^13,66-71^, are inversely related to PA participation. PA plays a pivotal role in the primary prevention and clinical treatment of many mental disorders ^72^ and is positively associated with mental health in pregnant women ^13^. Therefore, it is plausible that PA could exert beneficial intergenerational effects by preventing poor maternal mental health in the first place.

Mediators of the intergenerational effects of PA on offspring brain health and neurocognition may also be associated with preventing the ill-effects of poor maternal cardio-metabolic and sleep health. For example, PA is known to prevent diabetes ^14,73,74^, which is linked with adverse neurodevelopment including lower mental and psychomotor development in infants and lower intelligence quotient in school-aged children ^75-77^. Maternal hypertension ^78,79^ and obesity ^80^ are also recognised cardio-metabolic risk factors for impaired offspring neurodevelopment and can also be addressed by PA ^14,73,81-85^. Poor maternal sleep health during pregnancy has also been linked to aberrant offspring brain development and neurodevelopment ^86-88^ and can also be improved with PA participation ^89^.

Additionally, a critical direction for future human studies of PA will be to examine whether there could be similar and distinct mediating mechanisms of the intergenerational effects of PA between mothers (e.g., cord blood neurotrophin^90,91^ and neurosteroid^92,93^ concentrations, placental metabolism^94-96^), and fathers (e.g., sperm epigenetic programming)^97^.

### 4.4 Moderators

The majority of existing studies have focused on maternal PA during pregnancy (i.e., prenatal PA). There is still limited evidence in humans regarding the effects of preconception maternal PA on offspring brain health and neurocognition, and there is virtually no evidence examining the effects of paternal PA on offspring brain health and neurocognition; in fact, only one study in this review even reported paternal characteristics ^51^. In contrast, maternal and paternal PA (both prenatally and preconception) have been comparatively well-investigated in animal models ^33,97,98^. Another gap in our existing understanding of the intergenerational effects of parental PA on offspring brain health and neurocognition in humans is the timeframe in which any intergenerational effects of PA are strongest. The vast majority of studies in this review assessed brain health and neurocognition in the neonatal or infant stage, and did so using a variety of neurocognitive or developmental screening tests.

### 4.5 Strengths and limitations

Key limitations of this scoping review are that we only included studies published in the English language and only focused on elements of offspring brain and neurodevelopment that relate to neurocognition (i.e., we did not include studies that only reported on offspring motor development ^99,100^ or behaviour^101^). While the scoping review protocol was not prospectively registered and a formal assessment of methodological quality of the included studies was not performed, this is in accordance with guidelines for the conduct of scoping reviews ^34,35^. Other strengths of this scoping review are the comprehensiveness of the search, and that two investigators screened studies for eligibility and had “almost perfect agreement” ^102^.

## 5. CONCLUSION

Despite extensive evidence from animal models, little is known about the intergenerational effects of parental PA on offspring brain and neurocognitive development in humans, particularly paternal preconception PA. Evidence is promising but more experimental and large prospective observational studies with offspring follow-up into late adolescence and young adulthood are needed that have sophisticated strategies in place to promote participant retention and, for experimental studies, adherence to the PA intervention. Additionally, more objective and/or mechanistic assessments are also required as most existing evidence is based on subjective measures.

## Data Availability

All data produced in the present study are available upon reasonable request to the authors

## Notes

### Competing Interest Statement

The authors have declared no competing interest.

### Funding Statement

This study did not receive any funding

## REFERENCES

1. Taylor RM, et al. Effects of nutritional interventions during pregnancy on infant and child cognitive outcomes: A systematic review and meta-analysis. Nutrients. 2017;9(11):1265. https://doi.org/10.3390/nu9111265

2. Mattson SN, et al. Fetal alcohol spectrum disorders: a review of the neurobehavioral deficits associated with prenatal alcohol exposure. Alcoholism: Clinical and Experimental Research. 2019;43(6):1046–62. https://doi.org/10.1111/acer.14040

3. Lv S, et al. Association of maternal dietary patterns during gestation and offspring neurodevelopment. Nutrients. 2022;14(4):730. https://doi.org/10.3390/nu14040730

4. Han VX, et al. Maternal acute and chronic inflammation in pregnancy is associated with common neurodevelopmental disorders: a systematic review. Transl Psychiatry. 2021;11(1):71. https://doi.org/10.1038/s41398-021-01198-w

5. Cortés-Albornoz MC, et al. Maternal nutrition and neurodevelopment: A scoping review. Nutrients. 2021;13(10):3530. https://doi.org/10.3390/nu13103530

6. Chen H, et al. Neurodevelopmental effects of maternal folic acid supplementation: a systematic review and meta-analysis. Critical Reviews in Food Science and Nutrition. 2021:1–17. https://doi.org/10.1080/10408398.2021.1993781

7. Caut C, et al. Relationships between Women’s and Men’s Modifiable Preconception Risks and Health Behaviors and Maternal and Offspring Health Outcomes: An Umbrella Review. Semin Reprod Med. 2022. https://doi.org/10.1055/s-0042-1744257

8. Goodwin L, et al. Are psychosocial interventions effective in reducing alcohol consumption during pregnancy and motherhood? A systematic review and meta-analysis. Addiction (Abingdon, England). 2020. https://doi.org/10.1111/add.15296

9. Flynn A, et al. Dietary interventions in overweight and obese pregnant women: a systematic review of the content, delivery, and outcomes of randomized controlled trials. Nutr Rev. 2016;74(5):312–28. https://doi.org/10.1093/nutrit/nuw005

10. Chivu CM, et al. A systematic review of interventions to increase awareness, knowledge, and folic acid consumption before and during pregnancy. Am J Health Promot. 2008;22(4):237–45. https://doi.org/10.4278/06051566r2.1

11. Chamberlain C, et al. Psychosocial interventions for supporting women to stop smoking in pregnancy. Cochrane Database Syst Rev. 2017(2). https://doi.org/10.1002/14651858.cd001055.pub5

12. Davenport MH, et al. Impact of prenatal exercise on maternal harms, labour and delivery outcomes: a systematic review and meta-analysis. Br J Sports Med. 2019;53(2):99–107. https://doi.org/10.1136/bjsports-2018-099821

13. Davenport MH, et al. Impact of prenatal exercise on both prenatal and postnatal anxiety and depressive symptoms: a systematic review and meta-analysis. Br J Sports Med. 2018;52(21):1376–85. https://doi.org/10.1136/bjsports-2018-099697

14. Harrison CL, et al., editors. The role of physical activity in preconception, pregnancy and postpartum health. Semin Reprod Med; 2016: Thieme Medical Publishers. https://doi.org/10.1055/s-0036-1583530

15. Hayman M, et al. An investigation into the exercise behaviours of regionally based Australian pregnant women. Journal of Science and Medicine in Sport. 2016;19(8):664–8. https://doi.org/10.1016/j.jsams.2015.09.004

16. Vietheer A, et al. Sleep and physical activity from before conception to the end of pregnancy in healthy women: A longitudinal actigraphy study. Sleep Med. 2021;83:89–98. https://doi.org/10.1016/j.sleep.2021.04.028

17. American College of Obstetricians and Gynecologists. Physical Activity and Exercise During Pregnancy and the Postpartum Period: ACOG Committee Opinion, Number 804. 2020 Apr. Report No.: 0029-7844 Contract No.: 4. https://doi.org/10.1097/aog.0000000000003772

18. Australian Institute of Health and Welfare. Physical activity during pregnancy 2011–12. Canberra: AIHW; 2019. 978-1-76054-511-6 https://www.aihw.gov.au/reports/mothers-babies/physical-activity-during-pregnancy-2011-12

19. Brown WJ, et al. Australian guidelines for physical activity in pregnancy and postpartum. Journal of Science and Medicine in Sport. 2022. https://doi.org/10.1016/j.jsams.2022.03.008

20. Duncombe D, et al. Factors related to exercise over the course of pregnancy including women’s beliefs about the safety of exercise during pregnancy. Midwifery. 2009;25(4):430–8. https://doi.org/10.1016/j.midw.2007.03.002

21. Edvardsson K, et al. Giving offspring a healthy start: parents’ experiences of health promotion and lifestyle change during pregnancy and early parenthood. BMC Public Health. 2011;11(1):936. https://doi.org/10.1186/1471-2458-11-936

22. Brislane Á, et al. A Delphi Study to Identify Research Priorities Regarding Physical Activity, Sedentary Behavior and Sleep in Pregnancy. International journal of environmental research and public health. 2022;19(5):2909. https://doi.org/10.3390/ijerph19052909

23. Stillman CM, et al. Effects of exercise on brain and cognition across age groups and health states. Trends Neurosci. 2020. https://doi.org/10.1016/j.tins.2020.04.010

24. Valkenborghs SR, et al. The Impact of Physical Activity on Brain Structure and Function in Youth: A Systematic Review. Pediatrics. 2019;144(4):e20184032. https://doi.org/10.1542/peds.2018-4032

25. Erickson KI, et al. Physical activity, cognition, and brain outcomes: a review of the 2018 physical activity guidelines. Med Sci Sports Exerc. 2019;51(6):1242. https://doi.org/10.1249/mss.0000000000001936

26. Erickson KI, et al. Physical activity, fitness, and gray matter volume. Neurobiol Aging. 2014;35:S20–S8. https://doi.org/10.1016/j.neurobiolaging.2014.03.034

27. Valkenborghs SR, et al. Effect of high-intensity interval training on hippocampal metabolism in older adolescents. Psychophysiology. 2022:e14090. https://doi.org/10.1111/psyp.14090

28. Laurin D, et al. Physical activity and risk of cognitive impairment and dementia in elderly persons. Arch Neurol. 2001;58(3):498–504. https://doi.org/10.1001/archneur.58.3.498

29. Aberg MA, et al. Cardiovascular fitness is associated with cognition in young adulthood. Proc Natl Acad Sci U S A. 2009;106(49):20906–11. https://doi.org/10.1073/pnas.0905307106

30. Nyberg J, et al. Cardiovascular and cognitive fitness at age 18 and risk of early-onset dementia. Brain. 2014;137(5):1514–23. https://doi.org/10.1093/brain/awu041

31. Wilckens KA, et al. Exercise interventions preserve hippocampal volume: A meta-analysis. Hippocampus. 2021;31(3):335–47. https://doi.org/10.1002/hipo.23292

32. Yang Y, et al. Beneficial intergenerational effects of exercise on brain and cognition: a multilevel meta-analysis of mean and variance. Biol Rev Camb Philos Soc. 2021;96(4):1504–27. https://dx.doi.org/10.1111/brv.12712

33. Goli P, et al. Intergenerational influence of paternal physical activity on the offspring’s brain: A systematic review and meta-analysis. Int J Dev Neurosci. 2021;81(1):10–25. https://dx.doi.org/10.1002/jdn.10081

34. Peters MD, et al. Guidance for conducting systematic scoping reviews. Int J Evid Based Healthc. 2015;13(3):141–6. https://doi.org/10.1097/xeb.0000000000000050

35. Munn Z, et al. Systematic review or scoping review? Guidance for authors when choosing between a systematic or scoping review approach. BMC Med Res Methodol. 2018;18(1):143. https://doi.org/10.1186/s12874-018-0611-x

36. Covidence. Covidence systematic review software. Veritas Health Innovation Melbourne, Australia; 2019.

37. Cohen J. A coefficient of agreement for nominal scales. Educ Psychol Meas. 1960;20(1):37–46. https://doi.org/10.1177/001316446002000104

38. Norton K, et al. Position statement on physical activity and exercise intensity terminology. Journal of Science and Medicine in Sport. 2010;13(5):496–502. https://doi.org/10.1016/j.jsams.2009.09.008

39. Clapp JF, 3rd. Morphometric and neurodevelopmental outcome at age five years of the offspring of women who continued to exercise regularly throughout pregnancy. J Pediatr. 1996;129(6):856–63. https://doi.org/10.1016/s0022-3476(96)70029-x

40. Clapp JF, 3rd, et al. Neonatal behavioral profile of the offspring of women who continued to exercise regularly throughout pregnancy. Am J Obstet Gynecol. 1999;180(1 Pt 1):91–4. https://doi.org/10.1016/s0002-9378(99)70155-9

41. Clapp JF, 3rd, et al. The one-year morphometric and neurodevelopmental outcome of the offspring of women who continued to exercise regularly throughout pregnancy. Am J Obstet Gynecol. 1998;178(3):594–9. https://doi.org/10.1016/s0002-9378(98)70444-2

42. Domingues MR, et al. Physical activity during pregnancy and offspring neurodevelopment and IQ in the first 4 years of life. PLoS ONE Vol 9(10), 2014, ArtID e110050. 2014;9(10). https://dx.doi.org/10.1371/journal.pone.0110050

43. Ellingsen MS, et al. Neurodevelopmental outcome in 7-year-old children is not affected by exercise during pregnancy: follow up of a multicentre randomised controlled trial. BJOG. 2020;127(4):508–17. https://doi.org/10.1111/1471-0528.16024

44. Esteban-Cornejo I, et al. Maternal physical activity before and during the prenatal period and the offspring’s academic performance in youth. The UP&DOWN study. J Matern Fetal Neonatal Med. 2016;29(9):1414–20. https://dx.doi.org/10.3109/14767058.2015.1049525

45. Hellenes OM, et al. Regular moderate exercise during pregnancy does not have an adverse effect on the neurodevelopment of the child. Acta Paediatr. 2015;104(3):285–91. https://doi.org/10.1111/apa.12890

46. Jochumsen S, et al. Physical activity during pregnancy and intelligence in sons; A cohort study. Scand J Med Sci Sports. 2019;29(12):1988–95. https://dx.doi.org/10.1111/sms.13542

47. Jukic AMZ, et al. Physical activity during pregnancy and language development in the offspring. Paediatr Perinat Epidemiol. 2013;27(3):283–93. https://dx.doi.org/10.1111/ppe.12046

48. Labonte-Lemoyne E, et al. Exercise during pregnancy enhances cerebral maturation in the newborn: a randomized controlled trial. J Clin Exp Neuropsychol. 2017;39(4):347–54. https://doi.org/10.1080/13803395.2016.1227427

49. Menting MD, et al. Effects of maternal lifestyle interventions on child neurobehavioral development: Follow-up of randomized controlled trials. Scand J Psychol. 2019;60(6):548–58. https://dx.doi.org/10.1111/sjop.12575

50. Nakahara K, et al. Influence of physical activity before and during pregnancy on infant’s sleep and neurodevelopment at 1-year-old. Sci Rep. 2021;11(1):8099. https://dx.doi.org/10.1038/s41598-021-87612-1

51. Polanska K, et al. Maternal lifestyle during pregnancy and child psychomotor development - Polish Mother and Child Cohort study. Early Hum Dev. 2015;91(5):317–25. https://dx.doi.org/10.1016/j.earlhumdev.2015.03.002

52. Pryor J. Physical and behavioural correlates of 12-month development in small-for-gestational age and appropriately grown infants. J Reprod Infant Psychol. 1996;14(3):233–42. https://dx.doi.org/10.1080/02646839608404520

53. Mottola MF. Components of exercise prescription and pregnancy. Clin Obstet Gynecol. 2016;59(3):552–8. https://doi.org/10.1097/grf.0000000000000207

54. American College of Obstetricians and Gynecologists. ACOG committee opinion. Exercise during pregnancy and the postpartum period. Number 267, January 2002. 2002. Report No.: 0020-7292 Contract No.: 1. https://doi.org/10.1016/s0020-7292(02)80004-2

55. Hoffmann TC, et al. Better reporting of interventions: template for intervention description and replication (TIDieR) checklist and guide. Br Med J. 2014;348:g1687. https://doi.org/10.1136/bmj.g1687

56. Slade SC, et al. Consensus on Exercise Reporting Template (CERT): A Modified Delphi Study. Phys Ther. 2016. https://doi.org/10.2522/ptj.20150668

57. Teixeira R, et al. Completeness of retention data and determinants of attrition in birth cohorts of very preterm infants: a systematic review. Frontiers in Pediatrics. 2021;9:529733. https://doi.org/10.3389/fped.2021.529733

58. Costello L, et al. Informing retention in longitudinal cohort studies through a social marketing lens: Raine Study Generation 2 participants’ perspectives on benefits and barriers to participation. BMC Med Res Methodol. 2020;20(1):202. https://doi.org/10.1186/s12874-020-01074-z

59. Robinson KA, et al. Updated systematic review identifies substantial number of retention strategies: using more strategies retains more study participants. J Clin Epidemiol. 2015;68(12):1481–7. https://doi.org/10.1016/j.jclinepi.2015.04.013

60. Abshire M, et al. Participant retention practices in longitudinal clinical research studies with high retention rates. BMC Med Res Methodol. 2017;17(1):30. https://doi.org/10.1186/s12874-017-0310-z

61. Qiu A, et al. Prenatal maternal depression alters amygdala functional connectivity in 6 Month old infants. Transl Psychiatry. 2015;5(2). https://doi.org/10.1038/tp.2015.3

62. Dean DC, et al. Association of prenatal maternal depression and anxiety symptoms with infant white matter microstructure. Jama, Pediatr. 2018;172(10):973–81. https://doi.org/10.1001/jamapediatrics.2018.2132

63. Davis EP, et al. Prenatal maternal stress, child cortical thickness, and adolescent depressive symptoms. Child development. 2020;91(2):e432–e50. https://doi.org/10.1111/cdev.13252

64. Ortiz MT, et al. Maternal stress modifies the effect of exposure to lead during pregnancy and 24-month old children’s neurodevelopment. Environ Int. 2017;98:191–7. https://doi.org/10.1016/j.envint.2016.11.005

65. Polanska K, et al. Maternal stress during pregnancy and neurodevelopmental outcomes of children during the first 2 years of life. Journal of paediatrics and child health. 2017;53(3):263–70. https://doi.org/10.1111/jpc.13422

66. Kołomańska D, et al. Physical Activity and Depressive Disorders in Pregnant Women—A Systematic Review. Medicina. 2019;55(5):212. https://doi.org/10.3390/medicina55050212

67. Merced-Nieves FM, et al. Associations of prenatal maternal stress with measures of cognition in 7.5-month-old infants. Dev Psychobiol. 2021;63(5):960–72. https://doi.org/10.1002/dev.22059

68. Zhu P, et al. Does prenatal maternal stress impair cognitive development and alter temperament characteristics in toddlers with healthy birth outcomes? Dev Med Child Neurol. 2014;56(3):283–9. https://doi.org/10.1111/dmcn.12378

69. Ibanez G, et al. Effects of Antenatal Maternal Depression and Anxiety on Children’s Early Cognitive Development: A Prospective Cohort Study. PLoS One. 2015;10(8):e0135849. https://doi.org/10.1371/journal.pone.0135849

70. Lin Y, et al. Effects of prenatal and postnatal maternal emotional stress on toddlers’ cognitive and temperamental development. J Affect Disord. 2017;207:9–17. https://doi.org/10.1016/j.jad.2016.09.010

71. Urizar GG, Jr., et al. Role of Maternal Depression on Child Development: A Prospective Analysis from Pregnancy to Early Childhood. Child Psychiatry Hum Dev. 2022;53(3):502–14. https://doi.org/10.1007/s10578-021-01138-1

72. Biddle SJH, et al. Physical activity and mental health in children and adolescents: An updated review of reviews and an analysis of causality. Psychol Sport Exerc. 2019;42:146–55. https://doi.org/10.1016/j.psychsport.2018.08.011

73. Sanabria-Martínez G, et al. Effectiveness of physical activity interventions on preventing gestational diabetes mellitus and excessive maternal weight gain: a meta-analysis. BJOG. 2015;122(9):1167–74. https://doi.org/10.1111/1471-0528.13429

74. Aune D, et al. Physical activity and the risk of gestational diabetes mellitus: a systematic review and dose–response meta-analysis of epidemiological studies. Eur J Epidemiol. 2016;31(10):967–97. https://doi.org/10.1007/s10654-016-0176-0

75. Camprubi Robles M, et al. Maternal Diabetes and Cognitive Performance in the Offspring: A Systematic Review and Meta-Analysis. PLoS One. 2015;10(11):e0142583. https://doi.org/10.1371/journal.pone.0142583

76. Anderson JL, et al. Maternal obesity, gestational diabetes, and central nervous system birth defects. Epidemiology. 2005;16(1):87–92. https://doi.org/10.1097/01.ede.0000147122.97061.bb

77. Nomura Y, et al. Exposure to gestational diabetes mellitus and low socioeconomic status: effects on neurocognitive development and risk of attention-deficit/hyperactivity disorder in offspring. Arch Pediatr Adolesc Med. 2012;166(4):337–43. https://doi.org/10.1001/archpediatrics.2011.784

78. Grace T, et al. Maternal hypertensive diseases negatively affect offspring motor development. Pregnancy Hypertens. 2014;4(3):209–14. https://doi.org/10.1016/j.preghy.2014.04.003

79. Sun BZ, et al. Association of Preeclampsia in Term Births With Neurodevelopmental Disorders in Offspring. JAMA Psychiatry. 2020;77(8):823–9. https://doi.org/10.1001/jamapsychiatry.2020.0306

80. Tong L, et al. The impact of maternal obesity on childhood neurodevelopment. J Perinatol. 2021;41(5):928–39. https://doi.org/10.1038/s41372-020-00871-0

81. Cai C, et al. Prenatal Exercise and Cardiorespiratory Health and Fitness: A Meta-analysis. Med Sci Sports Exerc. 2020;52(7):1538–48. https://doi.org/10.1249/mss.0000000000002279

82. Perales M, et al. Benefits of aerobic or resistance training during pregnancy on maternal health and perinatal outcomes: A systematic review. Early Hum Dev. 2016;94:43–8. https://doi.org/10.1016/j.earlhumdev.2016.01.004

83. Aune D, et al. Physical activity and the risk of preeclampsia: a systematic review and meta-analysis. Epidemiology. 2014;25(3):331–43. https://doi.org/10.1097/ede.0000000000000036

84. Shiozaki A, et al. Risk factors for preeclampsia. Preeclampsia: Springer; 2018. p. 3–25. https://doi.org/10.1007/978-981-10-5891-2_1

85. Spracklen CN, et al. Physical Activity During Pregnancy and Subsequent Risk of Preeclampsia and Gestational Hypertension: A Case Control Study. Matern Child Health J. 2016;20(6):1193–202. https://doi.org/10.1007/s10995-016-1919-y

86. Nakahara K, et al. Association of maternal sleep before and during pregnancy with sleep and developmental problems in 1-year-old infants. Sci Rep. 2021;11(1):11834. https://doi.org/10.1038/s41598-021-91271-7

87. Peng Y, et al. Maternal sleep deprivation at different stages of pregnancy impairs the emotional and cognitive functions, and suppresses hippocampal long-term potentiation in the offspring rats. Mol Brain. 2016;9(1):17. https://doi.org/10.1186/s13041-016-0197-3

88. Nakahara K, et al. Association of physical activity and sleep habits during pregnancy with autistic spectrum disorder in 3-year-old infants. Commun Med (Lond). 2022;2:35. 10.1038/s43856-022-00101-y

89. Yang SY, et al. Effects of Exercise on Sleep Quality in Pregnant Women: A Systematic Review and Meta-analysis of Randomized Controlled Trials. Asian Nurs Res (Korean Soc Nurs Sci). 2020;14(1):1–10. https://doi.org/10.1016/j.anr.2020.01.003

90. Chouthai NS, et al. Changes in neurotrophin levels in umbilical cord blood from infants with different gestational ages and clinical conditions. Pediatr Res. 2003;53(6):965–9. https://doi.org/10.1203/01.Pdr.0000061588.39652.26

91. Ferrari N, et al. Exercise during pregnancy and its impact on mothers and offspring in humans and mice. J Dev Orig Health Dis. 2018;9(1):63–76. https://doi.org/10.1017/S2040174417000617

92. Aoyama B, et al. Role of neurosteroid allopregnanolone on age-related differences in exercise-induced hypoalgesia in rats. J Pharmacol Sci. 2019;139(2):77–83. https://doi.org/10.1016/j.jphs.2018.11.009

93. Crombie GK, et al. Neurosteroid-based intervention using Ganaxolone and Emapunil for improving stress-induced myelination deficits and neurobehavioural disorders. Psychoneuroendocrinology. 2021;133:105423. https://doi.org/10.1016/j.psyneuen.2021.105423

94. Ramírez-Vélez R, et al. Effect of exercise training on eNOS expression, NO production and oxygen metabolism in human placenta. PLoS One. 2013;8(11):e80225. https://doi.org/10.1371/journal.pone.0080225

95. Zhao SK, et al. Recreational physical activity before and during pregnancy and placental DNA methylation-an epigenome-wide association study. The American Journal of Clinical Nutrition. 2022. https://doi.org/10.1093/ajcn/nqac111

96. Chae SA, et al. Prenatal exercise in fetal development: a placental perspective. The FEBS Journal. 2022;289(11):3058–71. https://doi.org/10.1111/febs.16173

97. Vieira de Sousa Neto I, et al. Impact of paternal exercise on physiological systems in the offspring. Acta Physiologica. 2021;231(4):e13620. https://doi.org/10.1111/apha.13620

98. Fernandes J, et al. Physical exercise as an epigenetic modulator of brain plasticity and cognition. Neurosci Biobehav Rev. 2017;80:443–56. https://doi.org/10.1016/j.neubiorev.2017.06.012

99. Tinius R, et al. Maternal factors related to infant motor development at 4 months of age. Breastfeed Med. 2020;15(2):90–5. https://doi.org/10.1089/bfm.2019.0243

100. McMillan AG, et al. Effects of Aerobic Exercise during Pregnancy on 1-Month Infant Neuromotor Skills. Med Sci Sports Exerc. 2019;51(8):1671–6. https://doi.org/10.1249/mss.0000000000001958

101. Ji ES, et al. Effect of prenatal Qi exercise on mother–infant interaction and behavioral state. Journal of Child Health Care. 2014;19(4):504–12. https://doi.org/10.1177/1367493514522575

102. Landis JR, et al. The measurement of observer agreement for categorical data. Biometrics. 1977;33(1):159–74. https://doi.org/10.2307/2529310

