## Supplemental Information 1 for "The intergenerational effects of parental physical activity on offspring brain and neurocognition in humans: a scoping review"

**Supplemental Information 1. Search Terms**

**PUBMED**

(((((Pregnancy[Mesh:noexp] OR pregnancy[tiab] OR preconception[tiab] OR prenatal[tiab] OR maternal[tiab] OR paternal[tiab] OR intergenerational[tiab] OR transgenerational[tiab])

AND

(exercise[Mesh:noexp] OR muscle stretching exercises[mesh] OR physical conditioning, human[mesh] OR running[mesh] OR swimming[mesh] OR walking[mesh] OR exercise[tiab] OR physical activit*[tiab] OR running[tiab] OR swimming[tiab] OR walking[tiab] OR OR high-intensity interval training[tiab] OR resistance training[tiab] OR physical fitness[tiab] OR cardiorespiratory fitness[tiab] OR sedentary[tiab[ OR sitting time[tiab]))

AND

(offspring[tiab] OR infant[tiab] OR newborn[tiab] OR toddler[tiab] OR child*[tiab])

AND

(brain[tiab] OR hippocampus[Mesh:noexp] OR hippocampal volume[tiab] OR hippocampus[tiab] OR medial temporal lobe[tiab] OR prefrontal[tiab] OR brain structure[tiab] OR brain function[tiab] OR gr$y matter[tiab] OR white matter[tiab] OR connectivity[tiab] OR cognit*[tiab] OR executive function[tiab] OR memory [tiab] OR academic performance[tiab] OR intelligence[tiab] OR IQ[tiab])))

AND

humans[mesh])

NOT

(animals[mesh] OR mice[tiab] OR mouse[tiab] OR rat[tiab] OR rats[tiab] OR gerbils[tiab])))

**MEDLINE**

Pregnancy/ OR Prenatal Exposure Delayed Effects/ OR Maternal Behavior/ OR Maternal Exposure/ OR Paternal Behavior/ OR Paternal Exposure/ OR pregnancy.mp OR preconception.mp OR prenatal.mp OR maternal.mp OR paternal.mp OR intergenerational.mp OR transgenerational.mp

AND

Exercise/ OR Muscle Stretching Exercises/ OR Physical Conditioning, Human/ OR Physical Fitness/ OR Running/ OR Swimming/ OR Walking/ OR Physical Endurance/ OR High-Intensity Interval Training/ OR Resistance Training/ OR Oxygen Consumption/ OR Cardiorespiratory Fitness/ OR Muscle Strength/ OR Sedentary Behavior/ OR exercise.mp OR physical activit*.mp OR running.mp OR swimming.mp OR walking.mp OR physical endurance.mp OR high-intensity interval training.mp OR resistance training.mp OR physical fitness.mp OR cardiorespiratory fitness.mp OR muscle strength.mp OR sedentary.mp OR sitting time.mp

AND

Child/ OR Infant/ OR Infant, Newborn OR infant.mp OR newborn.mp OR toddler.mp OR child*.mp OR offspring.mp

AND

Brain/ OR Brain Cortical Thickness/ OR brain.mp OR Hippocampus OR Gray Matter/ OR White Matter/ OR Frontal Lobe/ OR Prefrontal Cortex/ OR Cognition/ OR Executive Function/ OR Memory/ OR Memory Consolidation/ OR Spatial Memory/ OR Memory, Short-Term/ OR Memory, Long-Term/ OR Memory, Episodic/ OR Academic Performance/ OR Educational Measurement/ OR Achievement/ OR Intelligence Tests/ OR Intelligence/ OR Child Development/ OR Infant Behavior/ OR Child Behavior/ OR Child Health/ OR Infant Health/ OR hippocampal volume.mp OR hippocampus.mp OR medial temporal lobe.mp OR prefrontal.mp OR brain structure.mp OR brain function.mp OR brain morphology.mp OR gr#y matter.mp OR white matter.mp OR connectivity.mp OR cognit*.mp OR neurocognition.mp OR executive function.mp OR memory.mp OR academic performance.mp OR intelligence.mp OR IQ.mp OR neurodevelopment.mp

AND

Humans/

**EMBASE**

Pregnancy/ OR Prenatal Exposure Delayed Effects/ OR Maternal Behavior/ OR Maternal Exposure/ OR Paternal Behavior/ OR Paternal Exposure/ OR pregnancy.mp OR preconception.mp OR prenatal.mp OR maternal.mp OR paternal.mp OR intergenerational.mp OR transgenerational.mp

AND

Exercise/ OR Muscle Stretching Exercises/ OR Physical Conditioning, Human/ OR Physical Fitness/ OR Running/ OR Swimming/ OR Walking/ OR Physical Endurance/ OR High-Intensity Interval Training/ OR Resistance Training/ OR Oxygen Consumption/ OR Cardiorespiratory Fitness/ OR Muscle Strength/ OR Sedentary Behavior/ OR exercise.mp OR physical activit*.mp OR running.mp OR swimming.mp OR walking.mp OR physical endurance.mp OR high-intensity interval training.mp OR resistance training.mp OR physical fitness.mp OR cardiorespiratory fitness.mp OR muscle strength.mp OR sedentary.mp OR sitting time.mp

AND

Child/ OR Infant/ OR Infant, Newborn OR infant.mp OR newborn.mp OR toddler.mp OR child*.mp OR offspring.mp

AND

Brain/ OR Brain Cortical Thickness/ OR brain.mp OR Hippocampus OR Gray Matter/ OR White Matter/ OR Frontal Lobe/ OR Prefrontal Cortex/ OR Cognition/ OR Executive Function/ OR Memory/ OR Memory Consolidation/ OR Spatial Memory/ OR Memory, Short-Term/ OR Memory, Long-Term/ OR Memory, Episodic/ OR Academic Performance/ OR Educational Measurement/ OR Achievement/ OR Intelligence Tests/ OR Intelligence/ OR Child Development/ OR Infant Behavior/ OR Child Behavior/ OR Child Health/ OR Infant Health/ OR hippocampal volume.mp OR hippocampus.mp OR medial temporal lobe.mp OR prefrontal.mp OR brain structure.mp OR brain function.mp OR brain morphology.mp OR gr#y matter.mp OR white matter.mp OR connectivity.mp OR cognit*.mp OR neurocognition.mp OR executive function.mp OR memory.mp OR academic performance.mp OR intelligence.mp OR IQ.mp OR neurodevelopment.mp

AND

Humans/

(NOTE: Limited search to English Language and Excluded Medline Journals)

**COCHRANE CENTRAL**

#1	pregnancy OR Prenatal OR Maternal OR Paternal OR preconception OR intergenerational OR transgenerational

#2	Exercise OR Muscle Stretching Exercises OR Physical Fitness OR Physical Conditioning Human OR Running OR Swimming OR Walking OR Physical Endurance OR High Intensity Interval Training OR Resistance Training OR Oxygen Consumption OR Cardiorespiratory Fitness OR Muscle Strength OR Sedentary Behavior OR physical activity OR physical fitness OR sitting time

#3	Child OR Infant OR newborn OR toddler OR offspring

#4	Brain OR Brain Cortical Thickness OR Hippocampus OR Gray Matter OR White Matter OR Frontal Lobe OR Prefrontal Cortex OR Cognition OR Executive Function OR Memory OR Memory Consolidation OR Spatial Memory OR Academic Performance OR Educational Measurement OR Achievement OR Intelligence Tests OR Intelligence OR Child Development OR Infant Behavior OR Child Behavior OR Child Health OR Infant Health OR hippocampal volume OR hippocampus OR medial temporal lobe OR prefrontal OR brain structure OR brain function OR brain morphology OR gr*y matter OR white matter OR connectivity OR cognit* OR neurocognition OR executive function OR memory OR academic performance OR intelligence OR IQ OR neurodevelopment

#5	#1 AND #2 AND #3 AND #4 in Trials

**PSYCHINFO**

Pregnancy/ OR Prenatal Exposure/ OR Transgenerational Patterns/ OR pregnancy.mp OR preconception.mp OR prenatal.mp OR maternal.mp OR paternal.mp OR intergenerational.mp OR transgenerational.mp

AND

Exercise/ OR Aerobic Exercise/ OR Conditioning/ OR Physical Fitness/ OR Sports/ OR Physical Activity/ OR Athletic Performance/ OR Physical Strength/ OR Running/ OR Swimming/ OR Walking/ OR Physical Endurance/ OR Training/ OR Athletic Training/ OR Obesity/ OR Body Mass Index/ OR Resistance/ OR Weightlifting/ OR Sedentary Behavior/ OR exercise.mp OR physical activit*.mp OR running.mp OR swimming.mp OR walking.mp OR physical endurance.mp OR high-intensity interval training.mp OR resistance training.mp OR physical fitness.mp OR cardiorespiratory fitness.mp OR muscle strength.mp OR sedentary.mp OR sitting time.mp

AND

Neonatal Period/ OR infant.mp OR newborn.mp OR toddler.mp OR child*.mp OR offspring.mp

AND

Brain/ OR Brain Connectivity/ OR Brain Size/ OR Hippocampus OR Gray Matter/ OR White Matter/ OR Frontal Lobe/ OR Prefrontal Cortex/ OR Cognition/ OR Cognitive Ability/ OR Executive Function/ OR Memory/ OR Memory Consolidation/ OR Spatial Memory/ OR Short Term Memory/ OR Long Term Memory/ OR Episodic Memory/ OR Academic Achievement/ OR Educational Measurement/ OR Achievement/ OR Intelligence Measures/ OR Intelligence/ OR Intelligence Quotient/ OR Childhood Development/ OR Cognitive Development/ OR Brain Development/ OR Neonatal Development/ OR Prenatal Development/ OR Infant Development/ OR Behavior/ OR Child Behavior/ OR Child Health/ OR Cognitive Ability/ OR brain.mp OR hippocampal volume.mp OR hippocampus.mp OR medial temporal lobe.mp OR prefrontal.mp OR brain structure.mp OR brain function.mp OR brain morphology.mp OR gr#y matter.mp OR white matter.mp OR connectivity.mp OR cognit*.mp OR neurocognition.mp OR executive function.mp OR memory.mp OR academic performance.mp OR intelligence.mp OR IQ.mp OR neurodevelopment.mp

AND

Humans/

(Note: Also limited to English Language)
