## Supplemental Table 1 for "The intergenerational effects of parental physical activity on offspring brain and neurocognition in humans: a scoping review"

*Supplemental Table 1. Summary of physical activity interventions*

| **First Author, Year** | **Frequency (sessions/week)** | **Intensity** | **Time**  **(min)** | **Type** | **Notes** |
| --- | --- | --- | --- | --- | --- |
| Ellingsen, 2020 | 3 | — | 45-60 | Aerobic, strength and balance | 12-week intervention between weeks 20-36 of gestation |
| Hellenes, 2014 | 1 | Moderate | — | — | Intervention delivered between weeks 20-36 of gestation |
| Labonte-Lemoyne, 2017 | 3 | Moderate | 20+ | — | — |
| Menting, 2019 |  |  |  |  |  |
| RADIEL study | 3-5 | Moderate-vigorous | 30-50 | — | Participants also aimed for >10,000 steps per day |
| LIFEstyle study | 2-3 | Moderate-vigorous | 30+ | — | Participants also aimed for >10,000 steps per day |
