## Supplemental Table 2 for "The intergenerational effects of parental physical activity on offspring brain and neurocognition in humans: a scoping review"

Supplemental Table 2. Summary of included studies

| **First Author, Year** | **Study Characteristics (Study design, N, country)** | **Parental Characteristics** | **Offspring Characteristics** | **Physical Activity Exposure (e.g., maternal vs. paternal, time, measure of physical activity)** | **Offspring Neurocognitive Outcome Assessed** | **Confounders Controlled for in Analyses** | **Key Findings** |
| --- | --- | --- | --- | --- | --- | --- | --- |
| ***Experimental*** | | | | | | | |
| Ellingsen, 2020 | Multi-site randomized controlled trial, n=875 (n=429 Exercise; n=426 Controls), sample decreased to n= 279 offspring (n=164 exercise; n=115 control) at follow-up, Norway  Hypothesis: No adverse effects of maternal physical activity on neurocognitive outcomes compared to controls. | Maternal Characteristics:  *Age (yr)*  Exercise: 30.5(3.9)  Control: 30.8(4.2)  *SES*  Exercise: 4.0(0.8)  Control: 4.1(0.8)  *BMI (kg/m^2^)*  Exercise: 24.3(2.8)  Control: 24.5(3.2)  *Exercise sessions per week* *at baseline:*  Exercise: 1.9(1.4)  Control: 1.7(1.4) | ***Sex (% male)***  **Exercise: 58.5**  **Control: 45.2**  *Gestational Age (weeks)*  Exercise: 39.9(1.7)  Control: 40.1(1.3)  *Birthweight (kg)*  Exercise: 3.5 (0.6)  Control: 3.6 (0.4)  *Head Circumference at Birth (*cm)  Exercise: 35.0 (2.1)  Control: 35.3 (1.4)  *Prematurity %*  Exercise: 4.3  Control: 2.6  *Admitted to NICU, %*  Exercise: 3.7  Control: 0.9 | *Treatment Group*: 12-week standardized exercise program between weeks 20-36 of pregnancy; Included aerobic, strength and balance exercises  3 sessions per week: 1 supervised group session lasting 60-minutes, and 2, 45-minute home sessions consisting of 30 minutes of choice aerobic activity and 15 minutes strength exercises.  *Control Group:* Standard antenatal care and education; Not discouraged from partaking in physical activity. | Parent report of motor skills, executive functions, perception, memory, language, learning, social skills, and emotional/ behavioural problems at 7 years (Five-to-Fifteen Questionnaire; FTF)  Relative risk (*Crude OR [95% CI])* of having more difficulties than peers:  *Motor Skills*  0.85 [0.39-1.84]  *Executive Functions*  1.11 [0.52- 2.38]  *Perception*  1.03 [0.49–2.16]  *Memory*  0.94 [0.42–2.13]  *Language*  0.88 [0.37–2.08] | Uncontrolled (crude) and adjusted models alternatively controlling for child age, child sex and maternal SES. | No significant differences in FTF scores at 7 years old between offspring of women in the exercise intervention group compared to those of women in the control condition.  Exercise during pregnancy has no adverse effects on offspring neurocognitive outcomes at 7 years. |
| Hellenes, 2014 | RCT, total n=336 (n=188 intervention; n=148 control), Norway  Hypothesis: Exercise during pregnancy would have no adverse effects on neurodevelopmental outcomes in offspring. | Maternal characteristics  *Age (yr)*  Exercise: 30.5 (4.1)  Control: 30.7 (3.9)  *SES*  Exercise: 3.9 (0.9)  Control: 4.0 (0.8)  *BMI (kg/m^2^)*  Exercise: 24.7 (3.1)  Control: 25.0 (3.7)  *Exercise sessions per week*  *(median [min, max])*  Exercise: 1.0 (0–5)  Control: 1.0 (0–5) | *Sex (% male)*  Exercise 55.3  Control: 45.3  *Gestational Age (wk)*  Exercise: 40.1 (1.2)  Control: 40.2 (1.2)  *Average Age at Assessment:*  Exercise: 20.1 months  Control: 20.1 months  *Birthweight (kg)*  Exercise: 3.5 (0.5)  Control: 3.6 (0.5)  *Head Circumference (cm)*  Exercise: 35.2 (1.7)  Control: 35.3 (1.6)  *Vaginal Delivery (n [%])*  Exercise: 142 (75.5)  Control: 108 (73.5) | *Intervention group:* Attended moderate intensity exercise group 1x per week from  weeks 20-36 of pregnancy.  *Control group:* physical activity levels/instructions (if any) were not reported. | BSID-III scores at 18-months (cognitive, language, and motor composites):  *Cognitive composite*  Exercise: 105.4 [103.8–106.9]  Control: 107.4 [105.7–109.2]  *Language composite*  Exercise: 98 [96.4–99.6]  Control: 98.5 [96.7–100.3]  *Motor composite*  Exercise: 98.6 [97.3–99.9]  Control: 100 [98.6–101.5]  18-month ASQ (communication, gross motor fine motor, problem solving, personal/social)  *Communication*  Exercise: 46 [44.0–48.1]  Control: 48.1 [45.7–50.4]  *Gross Motor*  Exercise: 57.6 [56.6–58.6]  Control: 57.7 [56.6–58.8]  *Fine motor*  Exercise: 57.3 [56.4–58.2]  Control: 56.9 [55.9–58.0]  *Problem Solving*  Exercise: 48.6 [47.1–50.1]  Control: 49.8 [48.1–51.4]  *Personal/Social*  Exercise: 51.8 [0.6–53.1]  Control: 53.1 [51.7–54.5] | Scores adjusted for:  gender, socio-economic  status, and number of siblings  ASQ scores were also adjusted for age  *Note: 34 statistical models considered but did not correct for multiple comparisons. | Moderate intensity exercise during pregnancy has no effect on the cognitive or language development of the offspring in early childhood. ITT analyses showed that motor skills also did not differ between the groups. |
| Labonte-Lemoyne, 2017 | Randomised controlled trial, n=18 (n=10 active v. n=8 sedentary), Canada  Hypothesis: Infant SPMMR will present a more mature  waveform in the children born to mothers who exercised during pregnancy. | Maternal Characteristics  *Age (yr)*  Active: 28.5 (3.4)  Sedentary: 27.8 (3.4)  *BMI (kg/m^2^)*  Active: 25.4 (2.0)  Sedentary: 25.3 (3.7)  *Pregnancy weight gain (kg)*  Active: 13.0 (3.0)  Sedentary: 14.9 (3.3)  *Pre-conception exercise habits (KPAS score)*  Active: 11.0 (1.7) Sedentary: 11.0 (1.2) | *Age at assessment (days)*  Active: 10.40 (1.90)  Sedentary: 9.75 (1.39)  *Weight (kg)*  Active: 3.4 (0.4)  Sedentary: 3.7 (0.4) | Active:  ≥20 min x 3 days per week @ RPE 12 (Borg 6-20 scale)  Sedentary:  Asked not to exercise | EEG (auditory oddball paradigm) acquired during active sleep for six ROIs (L+R x frontal, central, and parietal).  The newborns of mothers who exercised throughout their pregnancy have a smaller SPMMR than the children of mothers who remained sedentary (p=0.019). | N/A  Note:  There were no significant between-group differences on any demographic or socioeconomic measures assessed at entry to the study, but no matching reported. | Compared to newborns of mothers who were  inactive during their pregnancy, newborns of exercising pregnant women have more mature ERP amplitudes during a SPMMR task. |
| Menting, 2019 | Planned follow up of two RCTs of maternal lifestyle interventions before and during pregnancy, N = 258 total (n= 161 RADIEL, n = 96 LIFEstyle), Finland and Netherlands  Hypothesis: The benefits of the lifestyle interventions would be extended from the mothers to the children at ages 3 and 6 years. | *Maternal Characteristics (organized by RCT)* |  |  |  |  | Lifestyle interventions (which included a physical activity component) before or during pregnancy in obese women did not have an effect on child  neurobehavioral development. |
|  | n=161, Finland (RADIEL) | *Age (yr)*  Intervention: 33 (4)  Control: 32 (5)  *Pre-pregnancy BMI (kg/m^2^)*  Intervention: 35 (4)  Control: 34 (4) | *Sex (% male)*  Intervention: 52  Control: 49  *Gestational Age (weeks)*  Intervention: 40 (2)  Control: 40 (1)  *Age at follow up (yr)*  Intervention: 5.0 (0.3)  Control: 5.0 (0.3)  *Birthweight (kg)*  Intervention: 3.6 (0.6)  Control: 3.7 (0.5) | Intervention: 150 min moderate-vigorous intensity PA per week (either 30 min x 5 days or 50 min x 3 days) and ≥10,000 steps per day before and during pregnancy  Control: Diet and exercise education | *Neurodevelopment (ASQ) (median {IQR}, % abnormal)*  Intervention: 275 {253–290}, 0%  Control: 280 {255–290}, 6%  *Behavioural problems (Child Behavior Checklist) (median {IQR}, % abnormal)*  Intervention: 24 {11–3}, 4%  Control: 22 {12–32}, 6% | Child’s age, sex and conception method |  |
|  | n=96, Netherlands (LIFEstyle) | *Maternal characteristics*  *Age (yr)*  Intervention: 30 (4)  Control: 30 (4)  *Pre-pregnancy BMI (kg/m^2^)*  Intervention: 36 (3)  Control: 36 (3) | *Sex (% male)*  Intervention: 42  Control: 55  *Gestational Age (weeks)*  Intervention: 39 (2)  Control: 39 (2)  *Age at follow up (yr)*  Intervention: 4.1 (0.75)  Control: 4.3 (0.9)  *Birth weight (kg)*  Intervention: 3.4 (0.5)  Control: 3.5 (0.5) | Intervention: ≥30 min moderate-vigorous intensity PA x 2-3 days per week and ≥10,000 steps per day before and during pregnancy  Control: No PA intervention | *Neurodevelopment (Ages and stages questionnaire) (mean {IQR}, % abnormal)*  Intervention:  270 {255–285}, 3%  Control:  267 {244–281}, 0%  *Behavioural problems (Strengths and Difficulties Questionnaire) (mean {IQR}, % abnormal)*  Intervention:  8 {5–11}, 32%  Control:  8 {7–11}, 38% | Child’s age, sex, and conception method |  |
| ***Observational*** | | | | | | | |
| Clapp, 1996 | Prospective, case-control, observational design, n=40 (n=20 exercisers v. n=20 physically active controls), USA  Hypothesis: Continuing regular, vigorous, sustained exercise throughout pregnancy may adversely affect morphometric and neurodevelopmental outcomes in offspring at 5 years. | Maternal Characteristics  *Age (yr)*  Exercise: 31±1  Control: 31±1  *Body Fat (%)*  Exercise: 16.6±1.3  Control: 17.1±1.4  ***Pregnancy Weight Gain (kg)***  **Exercise: 12.8±1.1**  **Control: 16.2±1.4**  *VO_2max_ (mL/kg/min)*  Exercise: 54.4±2.6  Control: 53.1±2.9 | *Sex (% male)*  Exercise: 40  Control: 40  *Gestational Age (days)*  Exercise: 276±2  Control: 280±2 | Maternal (pre-conception and pre-natal)    *Running (n=11):*  21-68km per week @ 58-83% VO_2max_ (4.3-5.5 min per km).  *Aerobics (n=9):*  4-10 sessions per week (median=5) @ 62-88% VO_2max_ pre-conception | Neurodevelopmental characteristics at 5 years of age (Developmental Test Complex)  ***General intelligence* Exercise 125±2**  **Control: 117±3**  ***Oral language skill* Exercise: 119±2**  **Control: 109±3**  *Academic readiness* Exercise 100±2  Control: 98±2  ***Overall score***  **Exercise 115±l**  **Control: 111±2** | Parental groups were matched for smoking status, SES, education, marital stability, maternal and paternal morphometry, parity, maternal work outside the home, preconception fitness and exercise performance. Postnatal matching variables included lactation, type of childcare, parental exercise after birth, and maternal weight change since baseline.  Offspring were matched individually for sex, birth order, gestational age at delivery, clinically normal growth and development, absence of serious illness, type of childcare, and new siblings. | Overall neurodevelopmental scores were higher in the offspring of the exercising women due to differences in general intelligence and oral language skills.  No between-groups differences were seen in academic readiness skills, motor, or perceptual motor performance. |
| Clapp, 1998 | Prospective, case-control, observational design, n = 104 total expectant mothers and offspring (n=52 exercisers and n=52 matched controls), USA  Hypothesis: Continuing regular exercise throughout pregnancy  alters morphometric and neurodevelopmental outcomes of offspring. | Maternal Characterises  *Age (yr)*  Exercise: 31±1  Control: 31±1  ***Body Fat (%)***  **Exercise: 17.9±0.7**  **Control: 21.0 ±0.8**  ***Pregnancy Weight Gain (kg)***  **Exercise: 13.6±1.4**  **Control: 17.5±1.7**  *VO_2_ max (mL/kg/min)*  Exercise: 50.6±2.0  Control: 48.2±2.7 | *Sex (% male)*  Exercise: 46  Control: 56  *Gestational Age (days)*  Exercise: 277±1  Control: 279±1  ***Birthweight (kg)***  **Exercise: 3.38±0.06**  **Control: 3.48±0.07**  ***Body Fat (%)***  **Exercise: 9.5±0.4**  **Control: 12.6±0.6**  *Head Circumference (cm)*  Exercise: 34.6±0.2  Control: 34.8±0.3 | Maternal (pre-conception and pre-natal)    Exercising mothers: Ran (n=19), performed aerobics (n=23), or used one of several types of stair-climbing machines (n=10) throughout pregnancy 3+ times per week for 20+ minutes a session at an intensity averaging 75% of pre-pregnancy maximal aerobic capacity.  Controls: >60% occasional sustained LTPA during pregnancy. In the remainder of controls, performed LTPA ≤ 1x week in early and ≤ 1x every 2-3 weeks later in pregnancy. | BSID at 1 year old (mental and psychomotor scales only)  *Mental Score*  Exercise: 120±1  Control: 118±1  Mental Score Percentile  Exercise: 88±1  Control: 84±2  ***Psychomotor Score***  **Exercise: 108±1**  **Control: 101±2**  ***Psychomotor Score Percentile***  **Exercise: 69±3**  **Control: 53±4** | Exclusion criteria applied to eliminate known confounders in maternal sample.  Groups were approximately matched for age, body composition, health status, pregnancy complications, smoking, substance abuse, fitness, and family income, education, and home environment. | While offspring of the exercising mothers scored statistically higher on the Bayley psychomotor scale at 1 year, this result was not interpreted to be clinically meaningful.  There were no group differences in Bayley mental scores at 1 year. |
| Clapp, 1999 | Prospective observational, n = 65 total expectant mothers (n = 34 exercisers vs. n= 31 demographically similar inactive controls), USA  Hypothesis: Continuing regular exercise throughout pregnancy alters early neonatal behaviour. | Maternal Characteristics  *Age (yr)*  Exercise: 32±1  Control: 32±1  *Education (yr)*  Exercise: 16±1  Control: 16±1  *Pregestational Weight (kg)*  Exercise: 59.6±1.5  Control: 61.3±1.9  ***Weight Gain (kg)***  **Exercise: 12.8.±1.3**  **Control: 17.9±1.6**  *Active Labor >6 hr (%)*  Exercise: 26%  Control: 46% | *Sex (% male)*  Exercise: 53  Control: 48  *Gestational Age (days)*  Exercise: 279±1  Control: 278±1  ***Birthweight (kg)***  **Exercise: 3.44±0.06**  **Control: 3.64±0.05**  ***Body Fat (%)***  **Exercise: 9.7±0.4**  **Control: 13.1±0.5**  *Head Circumference (cm)*  Exercise: 34.6±0.2  Control: 34.7±0.4 | Maternal (preconception and prenatal) physical activity included running (n=11), aerobics (n=16), swimming (n=1), and stair machines (n=6) at >55-70% VO_2max_ for **≥**20min and **≥**3 times per week. | BNBAS at 5 days old  ***Orientation Behaviour***  **Exercise: 7.6±0.2**  **Control: 6.5±0.3**  ***State Regulation***  **Exercise: 6.6±0.2**  **Control: 4.9±0.3**  *Habituation (subsample n=34)*  Exercise: 8.1±0.2  Control: 8.1±0.1  Means and SEMs not reported for other Brazelton clusters. | Exercise and  control groups similar in age, education, pre-pregnancy weight, parity, birthing procedures, and Apgar scores. No explicit matching was reported. | Continuing regular  exercise throughout pregnancy altered early neurodevelopmental behaviour in a positive manner and had no detected negative effects.  Children from exercising women performed  better in 2 of the 6 domains of the Brazelton Scale 5 days after birth (orientation and  state regulation).  No differences detected in motor organization, autonomic stability, and range of state behaviours. |
| Domingues, 2014 | Prospective observational, n= 4231 newborns and moms, sample decreased to n= 3792 (48 months), Brazil  Hypothesis: Leisure Time Physical Activity (LTPA) during pregnancy can alter neurodevelopment of offspring during childhood. | Maternal Characteristics  *Age (*%)  ≥19 yr: 19%  20–35 yr: 68%  36+ yr: 13%  *Maternal Schooling (n (%))*  0–4 yr: 16%  5–8 yr: 41%  9–11 yr: 33%  12+ yr: 10% | 52 % male  14.6 % Preterm  9 % Low Birthweight | LTPA during pregnancy assessed retrospectively via questionnaire  *First trimester*  % Active: 10.6  0-149 min: 93.4%  150+ min: 6.6%  *Second trimester*  Active: 8.8 %  0–149 min: 95.1%  150+ min: 4.9%  *Third trimester*  Active: 6.7%  0–149: 96.5%  150+: 3.5% | Battelle’s Developmental Inventory P90 Score at 12  months of age and  IQ scores (Wechsler Preschool and Primary Scale of Intelligence). | Family income (quintiles), maternal schooling (four groups), smoking during pregnancy (yes/no), maternal age and skin colour (white / black / mixed), maternal occupation characteristics during pregnancy (standing for long periods and lifting heavy weights at work), number of previous births, maternal depressive symptoms at 48 months, birth weight, child’s sex, and preterm birth (yes, no). | LTPA during pregnancy  does not seem to negatively affect children’s neurodevelopment as  children from active mothers presented better results for most of the studied outcomes. After controlling for confounders, children from women who were active in the first, second and third trimester of pregnancy presented significant better results at 12 months of age. |
| Esteban-Cornejo, 2016 | Retrospective observational, n=1868, Spain  Objective To explore associations between maternal physical activity before and during pregnancy with academic performance in offspring. | Maternal Characteristics  ***University Educated (%)***  **Overall: 29**%  **Boy Offspring: 32**%  **Girl Offspring: 25**% | 50 % male  *Age (yr)*  Boys: 10.10 (3.30)  Girls: 10.26 (3.31)  ***Body Mass Index (kg/m2)***  **Boys: 19.42 (3.70)**  **Girls: 19.47 (3.66)**  ***Birthweight (kg)***  **Boys: 3.30 (0.56)**  **Girls: 3.17 (0.54)**  ***Gestational Age (wk)***  **Boys: 38.70 (2.39)**  **Girls: 38.70 (2.60)**  ***Fitness (z score)***  **Boys: 0.25 (1.01)**  **Girls: -.26 (0.75)**  ***Total PA (cpm)***  **Boys: 562.37(139.19)**  **Girls: 457.5 (122.43)** | Retrospective recall of physical activity levels before and during pregnancy, as well as changes in physical activity levels from pre to during pregnancy (yes/no).  *PA levels:*  *45% active before pregnancy*  *41% active during pregnancy*  *PA changes:*  30% remained Active  44% remained Inactive  11% became Active  15% became Inactive | Academic performance as measured by Math & Language grades and Grade Point Average at age 10  PA v. no PA (mean diff):  Preconception  Boys  **Math & Language: 0.43 [0.27, 0.59]**  **GPA: 0.33 [0.21, 0.45]**  Girls  Math & Language: 0.09 [-0.07, 0.25]  GPA: 0.09 [-0.03, 0.21]  Prenatal  Boys  **Math & Language: 0.28 [0.12, 0.44]**  **GPA: 0.19 [0.07, 0.32]**  Girls  Math & Language: 0.12 [-0.04, 0.28]  GPA: 0.12 [0.00, 0.24] | 3 models tested for each outcome:  *Model 1 covariates*:  Child age, birth city, maternal education  *Model 2 covariates:*  All from model 1 + gestational age, birthweight  *Model 3 covariates:*  All from model 2 + current BMI, physical activity, physical fitness | Maternal physical activity before and during pregnancy positively related to youth’s academic performance in boys, but not in girls.  Boys whose mothers were active before and during pregnancy had higher academic performance than those whose mothers continued being inactive or became active. |
| Jochumsen, 2019 | Retrospective observational study, n=4008 mother and son pairs, Denmark  Objective: To explore associations between regular maternal exercise during pregnancy and offspring intelligence. | *Maternal characteristics non-sport:*  *Age (%)*  <25 yr: 16%  25-29 yr: 43%  ≥30 yr: 41%  *BMI (kg/m^2^)(%)*  <18.5: 8%  18.5-24.99: 75%  ≥25: 17% | Offspring characteristics at 17-20 years, exercising mothers (‘non-sport’ moms)  100% male  *Gestational age at delivery (%)*  ≤37 wk: 43  37-39 wk: 51  ≥40 wk: 7  *Birth weight (%)*  <2.5 kg: 42  2.5-4.5 kg: 50  ≥4.5 kg: 7 | *Maternal physical activity in first trimester (Saltin‐Grimby Physical Activity Level Scale) (%)*  Sedentary: 43%  Light: 51%  Moderate-heavy: 7% | *Intelligence (Børge Priens test score) (adjusted mean difference)*  Sedentary: Reference  **Light: 1.1 [0.3, 1.8]**  **Moderate-heavy: 2.0 [0.7, 3.3]**  *Risk of low intelligence (adjusted odds ratio)*  Sedentary: Reference  **Light: 0.66 [0.49, 0.88]**  **Moderate-heavy: 0.46 [0.23, 0.93]** | Maternal body mass index, years in school, and smoking. | Higher levels of  physical activity and sport participation during pregnancy was associated with lower risk of low intelligence in early adulthood in males. |
|  |  | *Maternal characteristics sport:*  *Age (%)*  <25 yr: 17%  25-29 yr: 42%  ≥30 yr: 41%  *BMI (kg/m^2^)(%)*  <18.5: 8%  18.5-24.99: 74%  ≥25: 18% | Offspring characteristics at 17-20 years, (sport moms)  100% male  *Gestational age at delivery (%)*  ≤37 wk: 74  37-39 wk: 17  ≥40 wk: 9  *Birth weight (%)*  <2.5 kg: 73  2.5-4.5 kg: 17  ≥4.5 kg: 9 | *Maternal weekly sports participation in first trimester (%)*  0 hr: 74%  1-2 hr: 17%  3+ hr: 9% | *Intelligence (Børge Priens test score) (adjusted mean difference)*  0 hr: Reference  **1-2 hr: 1.5 [0.6, 2.5]**  **≥3 hr: 1.8 [0.6, 3.1]**  *Risk of low intelligence (adjusted odds ratio)*  0 hr: Reference  **1-2 hr: 0.50 [0.30, 0.83]**  ≥3 hr: 0.62 [0.35, 1.10] |  |  |
| Jukic, 2013 | Prospective observational study, n=7162 for primary analysis, USA  Hypothesis: Maternal physical activity during pregnancy will be associated with  more advanced language development in offspring. | *Maternal characteristics reported by time point* |  |  |  | Maternal education, crowding index, Home Observation for Measurement of Environment score, maternal age, race, head of household occupation, parity, anxiety, any infection in the first three months of pregnancy, weighted life events, depression, and hours worked during pregnancy, maternal parenting score, paternal parenting score, duration of breastfeeding, alcohol intake during pregnancy, smoking during pregnancy, fish consumption, sources of childcare, age at questionnaire completion |  |
|  | n=7162, 15 months follow up | *Age (%)*  <20 yr: 5%  21–25 yr: 21%  26–30 yr: 41%  31–35 yr: 25%  >35 yr: 8% | Not reported | *LTPA index (MET-h/week) at 18 weeks gestation (%)*  0: 13%  >0–4.4: 17%  4.5–15.1: 14%  15.2–18.1: 17%  18.2–27.6: 21%  ≥27.65: 17%  *General PA at 18 weeks gestation (hr/week) (%)*  0: 33%  0-1.9: 19%  2-3.9: 17%  4-5.9: 9%  6-10: 2%  >10: 10% | *MacArthur Infant*  *Communication scale*  *(Odds ratio of score >75th percentile, 95% CI) at 15 and 38 months*  LTPA index  0: Reference  >0–4.4: 0.97 [0.78, 1.21]  4.5–15.1: 1.17 [0.93, 1.47]  15.2–18.1: 1.08 [0.87, 1.35]  18.2–27.6: 1.16 [0.94, 1.43]  **≥27.65: 1.40 [1.13, 1.74]**  General PA (hr/week)  0: Reference  0-1.9: 1.01 [0.85, 1.20]  2-3.9: 1.09 [0.92, 1.31]  4-5.9: 1.19 [0.96, 1.46]  6-10: 1.06 [0.87, 1.29]  **>10: 1.26 [1.02, 1.55]** |  | Children of women in the highest quintile of prenatal LTPA and general PA were more likely to have better communication at 15-months. This effect was not present at 38 months. |
|  | n=4710, 8 years follow up | *Age (%)*  <20 yr: 3  21–25 yr: 18  26–30 yr: 42  31–35 yr: 28  >35 yr: 9 | Not reported | *LTPA index at 18 weeks gestation (%)*  0: 12%  >0–4.4: 17%  4.5–15.1: 15%  15.2–18.1: 16%  18.2–27.6: 21%  ≥27.65: 18%  *General PA at 18 weeks gestation (hr/week) (%)*  0: 33%  0-1.9: 20%  2-3.9: 17%  4-5.9: 9%  6-10: 11%  >10: 9% | *Verbal IQ (Wechsler Intelligence Scale) at 8 years (beta coefficient, 95% CI)*  LTPA index  0: Reference  >0–4.4: 1.12 [-0.48, 2.73]  4.5–15.1: 1.45 [-0.22, 3.11]  15.2–18.1: 0.45 [-1.17, 2.06]  18.2–27.6: 1.02 [-0.54, 2.58]  ≥27.65: 0.89 [-0.74, 2.53]  General PA (hr/week)  0: Reference  0-1.9: 0.55 [-0.69,1.79]  2-3.9: 0.003 [-1.30, 1.31]  4-5.9: -0.49 [-2.11, 1.13]  6-10: -1.40 [-2.94, 0.13]  **>10: -2.82 [-4.50, -1.14]** |  | Prenatal LTPA was not  associated with verbal IQ at 8 years of age, but higher prenatal general PA was associated with lower verbal IQ. |
| Nakahara, 2021 | Retrospective observational study, N = 74971 (prenatal) and N = 72700 (pregnancy), Japan  Hypothesis: Higher maternal PA before and during pregnancy may improve sleep and neurodevelopment in infants. | *Maternal characteristics* |  | International physical activity questionnaire | *Neurodevelopment at 1 year (ASQ)*  Adjusted relative risk of abnormal score for any 1 of 5 domains, reported by PA quartiles | Maternal age at delivery, smoking habits, alcohol consumption, pre-pregnancy BMI, parity, infertility treatment, type of delivery, gestational age at birth, infant sex, psychological distress at 1 year after delivery, Doctor diagnosis of asthma and atopic dermatitis at 1 year of age and feeding. | Higher levels of maternal PA, both before and during pregnancy, may reduce risk of developmental  problems in infants. |
|  | PA data before pregnancy: n=74971 | *Age (%)*  <25 yr: 9%  25–29 yr: 28%  30–34 yr: 36%  ≥35 yr: 28%  *BMI (kg/m^2^) (%)*  <18.5: 16%  18.5–24.9: 74%  ≥25.0: 10% | 51% male  *Gestational Age (%)* 37-38 wk: 33%  39-40 wk: 68%  *Birthweight (kg)*  3.1 (0.4) | *PA quartiles (METmin/week)*  Q_1_: 245 {168–385}  Q_2_: 791 {602–987}  Q_3_: 2163 {1617–2968}  Q_4_: 8078 {5789–12313}  n=12984: No PA | Q_1_: Reference  Q_2_: 0.95 [0.89, 1.00]  **Q_3_: 0.91 [0.86, 0.97]**  **Q_4_: 0.84 [0.79, 0.90]**  No PA: 1.05 [0.99, 1.11] |  |  |
|  | PA data during pregnancy: n=72700 | *Age (%)*  <25 yr: 9%  25–29 yr: 28%  30–34 yr: 36%  ≥35 yr: 27%  *BMI (kg/m^2^) (%)*  <18.5: 16%  18.5–24.9: 74%  ≥25.0: 10% | 51% male  *Gestational Age (%)* 37-38 wk: 33%  39-40 wk: 67%  *Birthweight (kg)*  3.1 (0.4) | During pregnancy  *PA quartiles (METmin/week)*  Q_1_: 196 {119-280}  Q_2_: 497 {399-595}  Q_3_: 1190 {987-1470}  Q_4_: 4774 {2968-7917}  n=16729 No PA | Q_1_: Reference  Q_2_: 0.96 [0.90, 1.02]  Q_3_: 0.95 [0.89, 1.01]  **Q_4_: 0.89 [0.84, 0.95]**  No PA: 1.09 [1.03, 1.16] |  |  |
| Polanska, 2015 | Prospective observational study, n=538 baseline (n = 424 @ 1year and n = 269 @ 2 years) mother–child pairs, Poland  Objective: To identify modifiable maternal lifestyle factors that impact child neurodevelopment. | Maternal characteristics  *Age (yr)*  30.5 (4.5)  *Pre-pregnancy BMI (kg/m^2^) (% )*  <18.5: 9%  18.5–24.9: 72%  25–29.9: 14%  ≥30: 5%  Paternal characteristics  *Age (yr)*  32.5 (5.5) | *Sex (% male)*  47  *Gestational Age (weeks)*  39.2 (1.4)  *Birthweight (kg)* 3.3 (0.5) | *Maternal LTPA (% participants)*  <3 METs or ≥3 METs and <2.5 h/week: 84%  ≥3 METs and ≥2.5 h/week: 16% | *Neurodevelopment (Bayley Scale)*  *≥3 METs and ≥2.5 h/week (β [95% CI])*  1-year-olds:  *Cognitive:*  −1.9 [−4.9, 1.1]  *Language:*  −1.4 [−4.4, 1.6]  *Motor*: −1.3 [−4.6, 1.9]  2-year-olds:  *Cognitive:*  0.4 [−3.4, 4.3]  ***Language:***  **4.1 [0.2, 8.0]**  *Motor*: 0.9 [−3.3, 5.0]  *<3 METs or ≥3 METs and <2.5 h/week:*  Reference | Examiner, parental age, parental education, child gender, and for  cognitive development additionally marital status and child day care attendance. | The recommended level of LTPA during pregnancy was beneficial for child language development at two years of age. |
| Pryor, 1996 | Longitudinal study, n=85 infants (n=65 small-for-gestational age infants; n=20 appropriate-grown infants), New Zealand  Objective: Initial identification of factors related to cognitive development in the first 2 years of life and which might be investigated further as potential predictors of infants who would benefit from early intervention. | Maternal characteristics  *Age (yr)* Small: 29.2 (6.1)  Appropriate: 31.0 (4.8) | ***Gestational Age (weeks)***  **Small: 39 (1)**  **Appropriate: 40 (1)**  ***Birthweight (kg)***  **Small: 2.6 (0.3)**  **Appropriate: 3.5 (0.3)** | *Prenatal exercise levels*  Small:  Little or no exercise: n=29  Moderate levels of exercise: n=29  High levels of exercise: n=7  Appropriate: Not reported  *Exercise in pregnancy (units not reported)*  Small: 1.7 (0.7) Appropriate: 1.7 (0.7) | Bayley Scales  of Infant Development (1 year of age)  *Mental Development Index*  Small: 95.2 (11.8)  Appropriate: 97.3 (12.9)  *Relationships with prenatal maternal exercise (correlation, variance)*  **Small: *r* = 0.52, 23%**  Appropriate: *r* = 0.04, n/a  *Infant Behaviour Record*  **Small: 62.7 (30.4)**  **Appropriate: 78.4 (25.6)** | Not reported | Prenatal maternal exercise levels were correlated with infant mental development at 12 months in small-for-gestational age infants, but not for appropriate-grown infants. |

Data reported as: mean (standard deviation); mean±standard error of the mean; mean [95% CI]; median{IQR}; counts/percentage. Bolded values were significantly different between groups.

Abbreviations: ASQ = Ages and Stages Questionnaire; BMI = body mass index; BSID = Bayley Scales for Infant Development; BNBAS = Brazelton Neonatal Behavioural Assessment Scales; EEG = electroencephalography; KPAS = Kaiser Physical Activity Survey; L = left; LTPA = leisure time physical activity; PA = physical activity; R = right; RCT = randomised controlled trial; ROI = region of interest; SES = socioeconomic status; SPMMR = slow positive mismatch response; yr = year;
